# Perceptions and use of traditional medicine/Centella Asiatica (Rau Ma) for cancer treatment in Vietnam: A quantitative survey study

**DOI:** 10.1101/2025.08.17.25333874

**Authors:** Yen Be Thi Hoang, Ky The Hoang, Huy Dinh Quoc, Michelle D. Garrett, Christopher J. Serpell, Rebecca Cassidy

## Abstract

This study investigates the public’s perceptions and practices regarding the use of *Centella asiatica* (*C. asiatica*), known locally as Rau Má, and potential as a cross-over drug for liver cancer, in Vietnam. The research employs a quantitative survey conducted across two major cities, Hanoi and Hue, to assess knowledge, attitudes, beliefs, and practices surrounding traditional medicine and its integration with modern healthcare, particularly in cancer treatment. A total of 804 participants were surveyed using a multistage stratified random sampling technique, ensuring a diverse representation across socio-economic backgrounds, ages, and urban/rural settings. Results reveal widespread awareness of *C. asiatica*, with significant socio-economic and regional differences in perceptions and acceptance of traditional remedies. Most participants, especially those from rural or lower socio-economic backgrounds, view treatment using derivatives of *C. asiatica* as a viable complementary therapy for liver cancer, with many expressing a preference for combining traditional medicine with conventional cancer therapies. The study highlights the potential for integrating traditional remedies like *C. asiatica* into modern healthcare practices and suggests further research to validate its efficacy and safety in cancer care.

## Introduction

Liver cancer is one of the leading causes of death in Vietnam, with the country ranking 7^th^ globally in terms of liver cancer incidence in 2022, with total of 24,502 new cases and 23,333 deaths from liver cancer. It is a critical public health concern [1]. In Vietnam, the integration of traditional and Western medicine varies, with traditional practices deeply rooted in the culture. A review discussing the integration of these medical systems noted that Vietnamese populations utilize both indigenous and foreign-influenced traditional medicines [2]. Traditional medicine, particularly the use of medicinal herbs, has long been a cornerstone of healthcare practices in many parts of the world, and use traditional medicine for treatment of liver cancer is receiving an increasing amount of attention [3]. In China, extensive investigations have been made in the traditional Chinese medicine for primary liver cancer over the past few years, especially in traditional Chinese medicine (TCM) treatment principles, improvement of therapeutic results, and prolonging of survival [4]. Recent studies have also highlighted the effect of traditional medicine on cancer-related fatigue reduction, while other studies have shown that herbs can have many beneficial effects in terms of health and quality of life improvement for cancer patients [5]. In Japan, traditional medicine is used in cancer-related palliative care (64.3%), with 32% of which used for related fatigue [6].

*C. asiatica*, commonly known as “Rau Ma,” has been the subject of numerous studies investigating its potential medicinal properties, particularly its anticancer effects. Several bioactive compounds in *C. asiatica*, including asiatic acid and madecassic acid and their glycosides, have demonstrated promising anticancer activities [7]. Madecassic acid, a triterpenoid compound found in *C. asiatica*, has been studied for its effects against liver cancer [8]. *C. asiatica* has been utilized in traditional medicine for treating various skin conditions, such as leprosy, lupus, varicose ulcers, eczema, and psoriasis [9]. Its efficacy is attributed to compounds like asiaticoside and madecassoside, which exhibit significant wound-healing activity through mechanisms including anti-inflammatory and antioxidative properties, collagen synthesis, and angiogenesis [10]. In Vietnam, the exploration of *C. asiatica*’s medicinal properties has led to collaborative research efforts. The UK-Vietnam C. asiatica Project (UV-CAP) has identified madecassic acid, a natural compound from *C. asiatica*, as active against liver cancer. Chemical derivatives of madecassic acid have shown further improved anticancer potential, underscoring the plant’s promise in cancer therapy [11].

This study seeks to investigate the knowledge, attitudes, beliefs, and practices of the Vietnamese public regarding the use of C. asiatica and other traditional medicines for liver cancer treatment. Through a quantitative survey in the cities of Hanoi and Hue, this research aims to gauge the level of awareness and acceptance of traditional medicine in general and *C. asiatica* in particular as a viable cancer treatment option. Understanding these factors is crucial in assessing the potential for integrating traditional medicines into mainstream medical practices and ensuring that patients have access to safe, effective treatment options.

## Methodology

### Study area

The study was conducted in two locations in Vietnam: Hanoi – the capital of Vietnam – and Hue (the central province). Both sides were selected to provide a broad and diverse representation of the Vietnamese population, with Hanoi and Hue each encompasssing both urban and rural areas, thus reflecting a range of socio-economic conditions.

The study was conducted across urban and rural districts in both cities, carefully selecting a high socio-economic district and a low socio-economic district within each city to ensure comprehensive data collection. In Hanoi, districts such as Phuc Tho (high socio-economic) and Thach That (low socio-economic) were selected, while in Hue, Huong Tra town (high socio-economic) and Quang Dien (low socio-economic) were chosen. By comparing these locations, the study was able to assess the impact of socio-economic status, education level, and urban *versus* rural location on the acceptance and practice of traditional medicine, particularly *C. asiatica*, for cancer treatment.

The differences between these cities and their respective socio-economic districts provided a unique opportunity to study the attitudes and practices regarding the use of traditional medicine across different settings in Vietnam. This diverse geographical and socio-economic approach enabled a more holistic understanding of the role that traditional medicines play in the treatment of serious diseases such as liver cancer.

### Study design and sampling

#### Study Design

The quantitative study employed a cross-sectional, descriptive survey design to investigate the knowledge, attitudes, beliefs, and practices related to the use of traditional medicine and *C. asiatica* for liver cancer treatment among the Vietnamese population. This design was chosen to provide a broad assessment of current behaviors and perceptions, offering a snapshot of public attitudes toward traditional medicine and its integration with modern cancer treatment. The survey incorporated both quantitative and descriptive elements, with structured questions aimed at assessing participants’ awareness of traditional medicine, their use of such treatments, and their views on the efficacy and safety of *C. asiatica* as an alternative or complementary treatment for liver cancer. Additionally, the study explored socio-demographic factors such as age, gender, education level, and income to better understand how these variables might influence attitudes toward traditional medicine.

#### Sampling Procedure

To ensure the sample was representative, the study used a multistage stratified random sampling technique, selecting participants from both urban and rural areas in Hanoi and Hue. In Hanoi, districts were classified as either high or low socio-economic areas based on income levels, healthcare access, and infrastructure development, with Phuc Tho District (urban) and Thach That District (peri-urban, lower-income) chosen for data collection. In Hue, Huong Tra town represented the urban, high socio-economic area, and Quang Dien District represented the rural, lower socio-economic area. We selected randomly 2 communes/wards from each target district for the survey. Selected communes are: Sen Phuong and Tho Loc (Phuc Tho district); Dai Dong and Lai Thuong (Thach That district); Tu Ha and Huong Van (Huong Tra town); and Quang An and Quang Tho (Quang Dien district). However, in Quang Dien District (Hue), the enumerator team was unable to interview the required number of participants in Quang Tho commune. As a result, the research team decided to randomly select Quang Phu commune of Quang Dien district, which is located next to Quang Tho commune and shares similar social and economic conditions, for inclusion in the survey. The originally allocated sample size for Quang Tho was 80 households, while for Quang Phu, it was 20 households..

The sample size for this study was calculated using Yamane Taro’s formula, designed to estimate the sample size for an unknown population [12]. The formula used was:

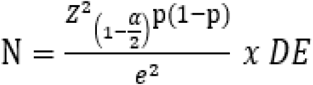

Where:

N = Desired sample size

z = value for selected alpha level (the standard 95% confidence interval will have alpha level of .025 in each tail, t = 1.96.

p = Proportion in the target population estimated. Normally, p=0.5 to ensure that accumulation of p(1-p) is highest. This will ensure the estimated samples.

e = Marginal of error (+-0.05 (5%) of error is acceptable)

DE (Design effect): because simple random sampling is not applied, the sample size should multiply with the design effect. DE = 2 is used in this survey

The aim was to obtain a sample size that would ensure a 95% confidence level, a 5% margin of error, and a design effect (DE) of 2. Based on these parameters, the calculated sample size for the target population was 768 respondents. Accounting for an estimated 5% rate of incomplete questionnaires, the required total sample size was adjusted to 800, after rounding down. In practice, a total of 804 respondents were successfully surveyed during fieldwork.

Within each district, households were randomly selected, and one adult (18 years or older) from each household participated, ensuring a representative and random sample. An additional 10% of participants were recruited to account for non-responses and incomplete data, ensuring the target sample size of 800 respondents, with a balanced mix of males and females (Table 1). The sample was stratified into three socio-economic status groups—low, middle, and high—allowing for analysis of how attitudes toward traditional medicine, including *C. asiatica*, varied across different socio-economic backgrounds. Data collection occurred over four weeks with trained enumerators conducting face-to-face interviews, asking participants about their socio-demographics, health conditions, and views on *C. asiatica* and traditional medicines.

**Table 1.** Size of Innitial Planned Sample Selection and Corresponding Sample Sizes Province Hanoi (400) Hue (400)

| Province | Hanoi (400) |  | Hue (400) |  |
| --- | --- | --- | --- | --- |
| District | High socio-economic (Phuc Tho) | Low socio-economic (Thach That) | High socio-economic (Huong Tra town) | Low socio-economic (Quang Dien) |
| Sample size | 200 | 200 | 200 | 200 |

### Ethical Considerations

This study complies with local and international ethical requirements. The study protocol (Application Ref: SRCEA0280) was approved by the ethics committee of the University of Kent on November 20^th^ 2020, as the part of the research project Homegrown solutions in Viet Nam: bringing *C. asiatica* compounds forwards as candidates to treat liver cancer, WP6: Laying foundations for the future.

Ethical approval for the study was shared with the local relevant institutions in Hanoi and Hue who then supported conducting the survey in target areas. Before administering each questionnaire, each targeted person was explained what the study concerned, and free and informed consent was obtained from all participants, who were assured of the confidentiality of their responses. Participation in the study was voluntary, and participants were free to withdraw at any time without penalty. Consent was confirmed by signing the form prepared for this purpose. All survey participants were given the study information leaflet (PIL) in Vietnamese or English, and could ask any questions before deciding whether to take part. Consent forms were signed by all participants. No minors were involved in the study

### Data collection

Data for this study was collected from 1^st^ to 29^th^ April 2022 through a structured questionnaire administered by trained enumerators in face-to-face interviews. The use of face-to-face interviews was chosen to enhance response accuracy and ensure clarity of questions and answers, thereby minimizing misunderstandings and potential misinterpretation that might occur with self-administered surveys. The questionnaire was designed to assess various aspects of participants’ knowledge, attitudes, beliefs, and practices related to the use of traditional medicines/*C. asiatica* for liver cancer treatment. It included both closed-ended and Likert-scale questions, which allowed respondents to express the extent to which they agreed or disagreed with specific statements about their use of traditional medicine, their perceptions of its safety and efficacy, and the likelihood of integrating it with conventional cancer treatments.

The survey was administered to participants across two major cities, Hanoi and Hue, ensuring diverse representation from both urban and rural areas. Trained enumerators, chosen for their cultural and linguistic familiarity with the local population, conducted the interviews. They underwent a rigorous training program to familiarize themselves with the survey’s objectives, ethical considerations, and the procedures for obtaining informed consent from respondents. Prior to conducting the survey, each participant was provided with an informed consent form, outlining the study’s purpose, potential risks, and the voluntary nature of participation, assuring them of the confidentiality of their responses.

The survey covered several key sections: demographic information (age, gender, socio-economic status, and education level), health and cancer history, knowledge about traditional medicine, attitudes toward the use of *C. asiatica* in cancer treatment, and respondents’ willingness to use or recommend it [13]. The interviews typically lasted between 15 to 30 minutes. To test the effectiveness of the survey instrument and ensure clarity, a pilot study was conducted with a small sample of 30 participants, leading to minor adjustments in the final questionnaire.

### Statistical analysis

Data analysis for this study was conducted using a combination of descriptive statistics, inferential statistics, and regression analysis to examine the relationship between participants’ demographic characteristics and their attitudes and behaviors regarding traditional medicine use. The data analysis process was carried out in several steps, as detailed below:

#### Data Cleaning

Data analysis for this study involved both descriptive and inferential statistical techniques to assess the knowledge, attitudes, and practices regarding traditional medicine/*C. asiatica* and its use for cancer treatment First, the raw data was cleaned to ensure accuracy. This included handling missing values and identifying potential outliers, which were addressed appropriately. Descriptive statistics were used to summarize the demographic characteristics of the study population and key findings. Frequencies and percentages were computed for categorical variables such as gender, education level, and location, while means and standard deviations were calculated for continuous variables, including age and Likert-scale responses.

Inferential statistics were then applied to examine associations between demographic factors and attitudes toward traditional medicine. Chi-square tests were used to assess relationships between categorical variables, such as the association between socio-economic status and knowledge of traditional medicines/*C. asiatica*, and to explore patterns in participants’ willingness to use traditional medicines/*C. asiatica* for liver cancer treatment. For continuous variables, such as the level of agreement with specific statements about traditional medicine, independent t-tests were used to compare two groups (e.g., urban *vs*. rural respondents), and analysis of variance (ANOVA) was applied when comparing three or more groups, such as different age groups or education levels.

To further understand the factors influencing the willingness to incorporate traditional medicines/*C. asiatica* into cancer treatment, logistic regression analysis was conducted. This method allowed for the examination of multiple independent variables (such as socio-economic status, age, education level, and previous use of traditional medicine) to determine which factors were statistically significant predictors of participants’ likelihood to use or recommend traditional medicine/*C. asiatica*. All statistical analyses were performed using Stata v14.2, a widely used software tool in social science and health research.

## Results

### Survey population

The study successfully surveyed a total of 804 respondents from Hanoi and Hue, ensuring diversity across socio-economic statuses, gender, and age groups. The sample was designed to be representative of both urban and rural populations, allowing for a comprehensive analysis of attitudes toward traditional medicine in different contexts.

Table 2 presents the demographic characteristics of the study population, disaggregated by socio-economic status. The proportion of adults participating in the study was evenly distributed between locations (Hanoi and Hue), genders (male and female), and 03 age groups (under 40, 40-60, over 60 in general and by socioeconomic status in each region. In terms of education level, the majority of people has an education level of either high school or lower: lower secondary school (~39%), high school: 23.3%, and primary school (21.5%); and, there was a difference between that of people in the higher socio-economic group and those of lower socioeconomic status (χ^2^ test, p < 0.001). The proportion of survey participants who were heads of households was 61.2% and was higher in the high socioeconomic group (64.7%) compared to that of the lower socioeconomic group (χ^2^ test, p=0.04). The main source of income of the participatants was agriculture (51.1%) and waged employment (30.7%). The proportion of people whose income came from agriculture was higher in areas with low socioeconomic areas and, in contrast, the percentage of people whose income came from waged employment was higher in the high socio-economic areas (p< 0.001).

**Table 2.** Demographic Information of the Study Population.

| Indicators | Low socio-economic (n=402) | High socio-economic (n=402) | Overall (n=804) | P value |
| --- | --- | --- | --- | --- |
| Location (%) |  |  |  |  |
| *Hanoi* | 50.2 | 50.0 | 50.1 | 0.94 |
| *Hue* | 49.8 | 50.0 | 49.9 |  |
| Gender (%) |  |  |  |  |
| *Male* | 49.8 | 49.8 | 49.8 | 1.0 |
| *Female* | 50.2 | 50.2 | 50.2 |  |
| Age (mean ± SD) | 54.4 ± 11.9 | 54.3 ± 11.3 | 54.4 ± 11.6 | 0.89 |
| Age group (%) |  |  |  |  |
| *Under 40 yrs* | 14.7 | 13.7 | 14.2 | 0.92 |
| *40-59 yrs* | 36.1 | 36.3 | 36.2 |  |
| *60+ yrs* | 49.2 | 50.0 | 49.6 |  |
| Educational level (%) |  |  |  |  |
| *Primary and less* | 22.9 | 20.2 | 21.5 | <0.001 |
| *Secondary* | 43.8 | 33.3 | 38.5 |  |
| *High school* | 21.1 | 25.4 | 23.3 |  |
| *High education* | 12.2 | 21.1 | 16.7 |  |
| Head of the household (%) | 64.7 | 57.7 | 61.2 | 0.04 |
| Income source (%) |  |  |  |  |
| *Waged employment* | 23.4 | 38.1 | 30.7 | <0.001 |
| *Farmers* | 60.2 | 42.0 | 51.1 |  |
| *Small traders* | 11.7 | 16.4 | 14.1 |  |
| *Others (freelancers etc.)* | 4.7 | 3.5 | 4.1 |  |

The majority of surveyed households have Household Wealthy Index [14] belonging to either the rich (~36%) or the richest (~58%) levels. The proportion of richest households in high socioeconomic areas were higher than that of the low socioeconomic areas (64.7% vs 50.8%, χ^2^ test, p<0.001).

Table 3 shows the health conditions by age group and socio-economic status reveals important patterns and significant differences from the survey. Overall, 27.9% of respondents use long-term medication, with higher usage among older age groups, peaking at 32.3% for those aged 60+. Statistically significant differences were observed between socio-economic groups, with 22.9% in low socio-economic areas and 24.3% in high socio-economic areas using long-term medication (p<0.05).

**Table 3.**
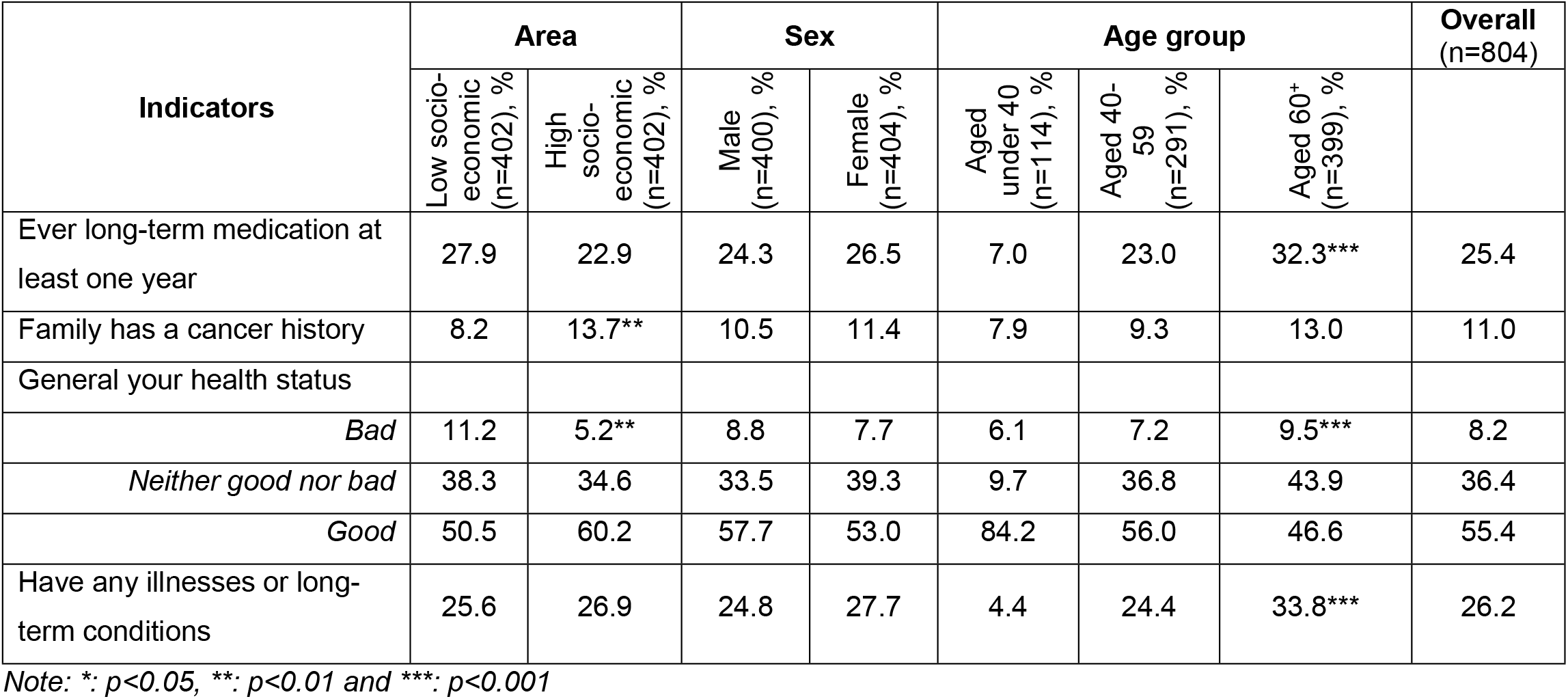
Health condition of surveyed respondents by area, sex and age group.

Regarding family cancer history, 8.2% of respondents reported a family history of cancer, with a higher prevalence in the high socio-economic group (13.7%) compared to the low socio-economic group (10.5%) (p<0.01). Family cancer history increases with age, reaching 13.0% in those aged 60+.

Health status declines with age, as 84.2% of those under 40 report good health, compared to only 46.6% in those aged 60+ (p<0.001). Statistically significant differences were also found in socio-economic groups, with 60.2% of low socio-economic respondents reporting good health compared to 50.5% in the high socio-economic group (p<0.01).

The prevalence of long-term illnesses rises with age, from 4.4% in those under 40 to 33.8% in those aged 60+ (p<0.001). While no significant difference was found between socio-economic groups, the data highlights the growing burden of chronic conditions with age.

In summary, both age and socio-economic status significantly affect health outcomes, with older individuals experiencing more health issues and higher reliance on long-term medication. Socio-economic disparities also influence general health status and chronic conditions.

### Knowledge of *C. asiatica* and its Use

The study revealed that a large proportion of respondents (87.3%) were familiar with *C. asiatica*, reflecting its widespread recognition as a traditional remedy (Figure 1). However, there were notable differences in the level of awareness depending on the socio-economic background and education levels of the participants. Specifically, respondents from higher socio-economic areas demonstrated a higher level of awareness and understanding of the medicinal properties of *C. asiatica* compared to their counterparts in lower socio-economic regions.

**Fig 1.**
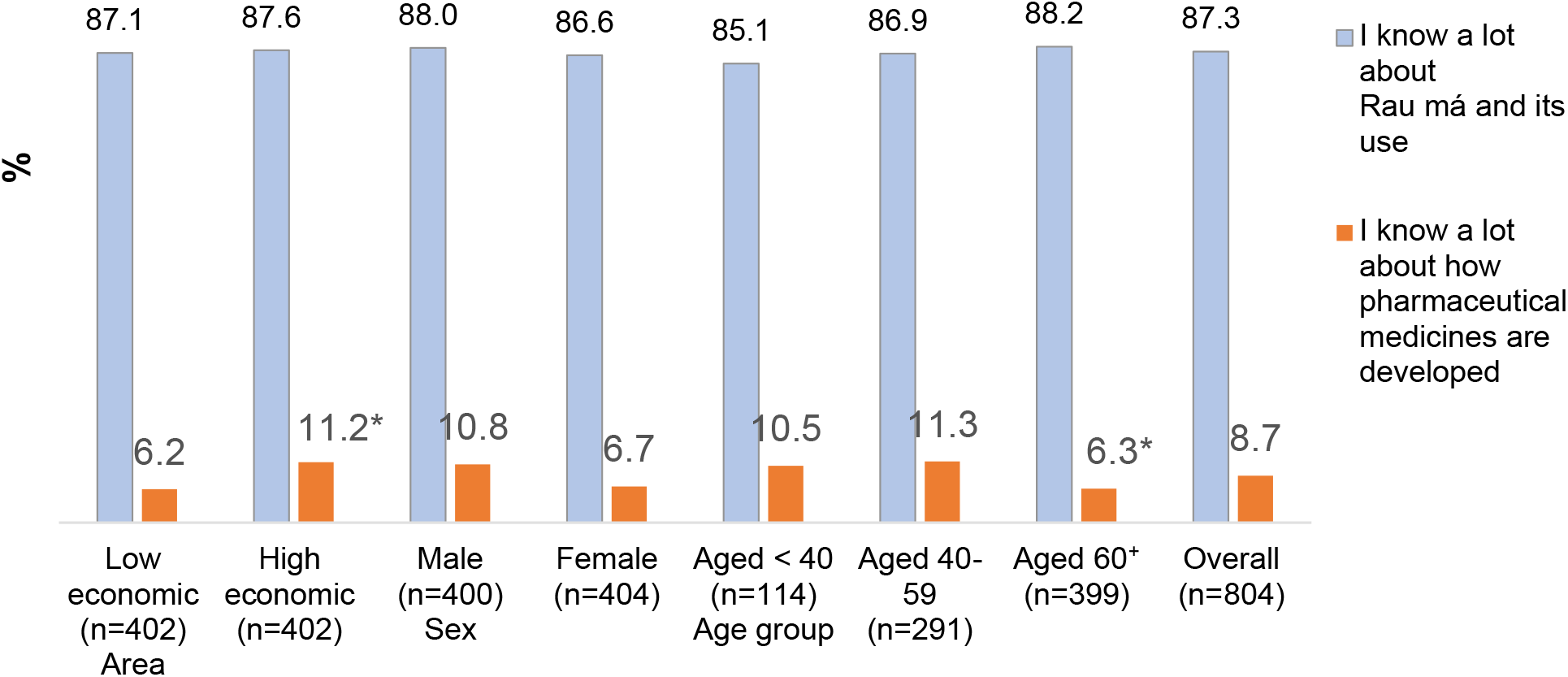
Knowledge of medical properties of *C. asiatica* by Socio-Economic Status, sex and age group.

Figure 2 illustrates the proportion of respondents who recognized *Centella asiatica* as having medicinal properties across different socio-economic areas, age groups, and sexes. In high socio-economic areas, nearly 68% of respondents associated *C. asiatica* with medicinal properties, reflecting a more developed understanding of its potential health benefits. These respondents were generally more exposed to healthcare information, whether through formal education, healthcare systems, or media channels, which may have contributed to their higher awareness. In contrast, 55% of respondents from low socio-economic areas identified *C. asiatica* as having medicinal value, indicating a significant knowledge gap between the two groups (~68% vs 55%, χ^2^ test, p< 0.001). This disparity is likely linked to differences in education levels, access to healthcare resources, and exposure to traditional medicine practices.

**Fig 2.**
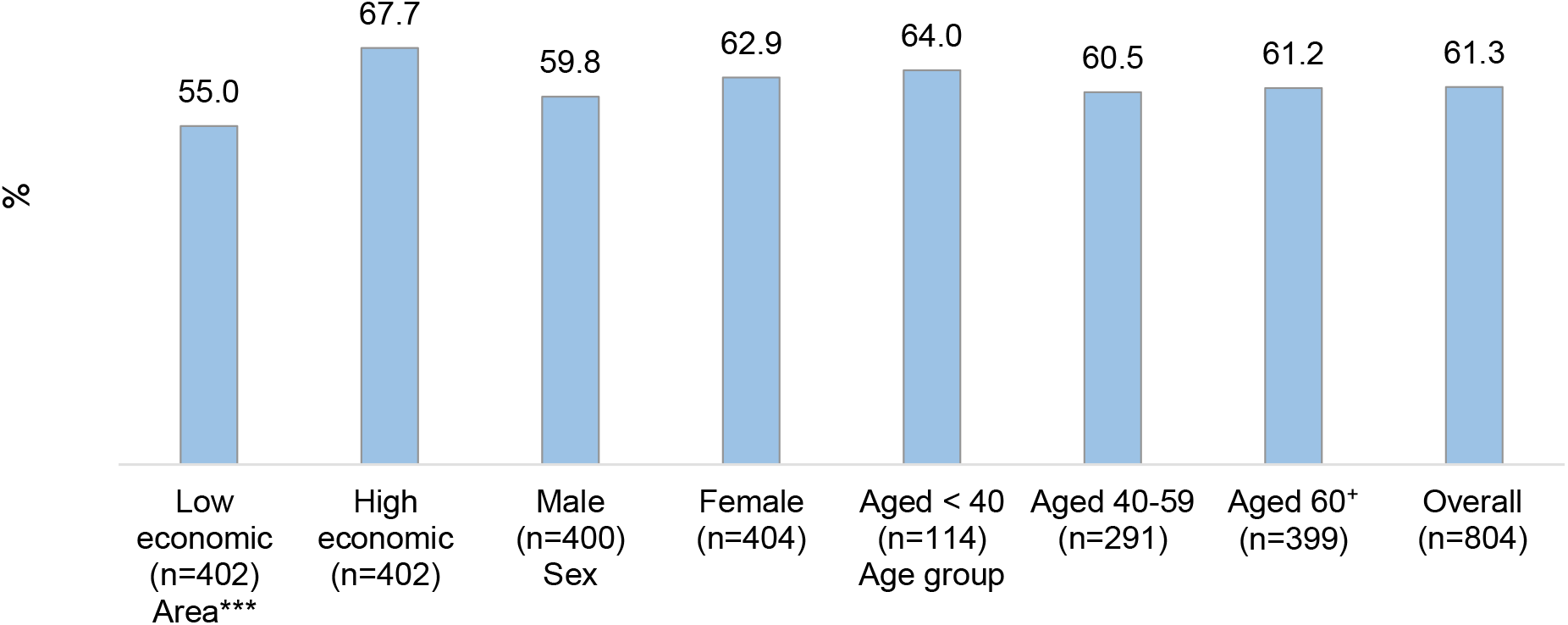
Respondents reported centella asiatica has medical properties by area, sex and age group.

### Attitudes Toward Traditional Medicine

Our study revealed interesting insights into respondents’ attitudes toward traditional medicine compared to Western medicine. A significant portion of respondents (74.4%) agreed that the development of new medications is essential, showing strong support for innovation in the field of modern medicine. However, an even more noteworthy finding was that 67.8% of respondents believed that traditional medicine was safer than pharmaceutical treatments. This suggests that, despite the availability of modern treatments, traditional remedies continue to be regarded favorably for their perceived safety and fewer side effects.

### Safety and Efficacy of Traditional Medicine

Table 5 presents the distribution of respondents’ opinions on traditional medicine, disaggregated by socio-economic area, gender, and age group. The proportion of respondents who agreed that traditional remedies, such as Centella asiatica to be safer than western medicine was nearly 68%. This figure was 16% higher in the low socio-economic areas compared to that of the other group (75.6% vs. 60%, χ^2^ test, p<0.001). It also supports the idea that many people in Vietnam, especially those from rural areas or lower socio-economic backgrounds, feel more comfortable with the “natural” and holistic approach that traditional medicines offer. Additionally, while only approximately one-third of respondents (30–36%) agreed that herbal remedies are more effective than pharmaceutical drugs, this belief was more pronounced among individuals aged 40 to 59, reflecting generational variations in medical trust and knowledge. The perceived synergistic potential of combining herbal and Western medicines—endorsed by around 40% of participants—further emphasizes the integrative approach favored by many patients. Collectively, these findings highlight the enduring role of traditional medicine as a complementary or alternative option

**Table 5.** Opinions on traditional medicine of survey participants by region, gender and age group.

| Indicator | Area |  | Sex |  | Age group |  |  | Overall<br>(n=804) |
| --- | --- | --- | --- | --- | --- | --- | --- | --- |
|  | Low socio-economic<br>(n=402),<br>% | High socio-economic<br>(n=402),<br>% | Male<br>(n=400),<br>% | Female<br>(n=404),<br>% | Aged under 40<br>(n=114),<br>% | Aged 40-59<br>(n=291),<br>% | Aged 60-70<br>(n=399),<br>% |  |
| Herbal remedies/Vietnamese medicine are more effective than [pharmaceutical] treatments | *** |  |  |  |  |  |  |  |
| *Disagree* | 17.9 | 46.0 | 32.3 | 31.7 | 31.6 | 34.7 | 30.1 | 32.0 |
| *Neutral* | 47.3 | 23.6 | 35.2 | 35.6 | 45.6 | 28.9 | 37.3 | 35.4 |
| *Agree* | 34.8 | 30.4 | 32.5 | 32.7 | 22.8 | 36.4 | 32.6 | 32.6 |
| Herbal remedies are safer than [pharmaceutical] | *** |  |  |  |  |  |  |  |
| *Disagree* | 6.2 | 25.1 | 15.8 | 15.6 | 16.7 | 15.8 | 15.3 | 15.7 |
| *Neutral* | 18.2 | 14.9 | 19.0 | 14.1 | 23.7 | 13.4 | 16.8 | 16.5 |
| *Agree* | 75.6 | 60.0 | 65.2 | 70.3 | 59.6 | 70.8 | 67.9 | 67.8 |
| Herbal remedies only work because of the placebo effect | *** |  |  |  |  |  |  |  |
| *Disagree* | 48.5 | 61.0 | 53.2 | 56.2 | 54.4 | 55.3 | 54.4 | 54.7 |
| *Neutral* | 32.6 | 30.3 | 32.8 | 30.2 | 35.1 | 27.8 | 33.1 | 31.5 |
| *Agree* | 18.9 | 8.7 | 14.0 | 13.6 | 10.5 | 16.9 | 12.5 | 13.8 |
| Herbal medicine works well in combination with western medicine | *** |  |  |  |  |  |  |  |
| *Disagree* | 25.6 | 14.7 | 19.5 | 20.8 | 17.5 | 21.6 | 19.8 | 20.2 |
| *Neutral* | 37.8 | 41.0 | 42.0 | 36.9 | 50.0 | 36.8 | 38.4 | 39.4 |
| *Agree* | 36.6 | 44.3 | 38.5 | 42.3 | 32.5 | 41.6 | 41.8 | 40.4 |

Note: \*: $p < 0.05$ , \*\*: $p < 0.01$ and \*\*\*: $p < 0.001$

The majority do not agree that herbal remedies work solely because of the placebo effect. Specifically, approximately 54.7% of respondents disagreed with this statement, whereas only about 13.8% agreed. This indicates that most people believe in the actual pharmacological efficacy of herbal medicines rather than attributing their effects merely to psychological factors. When segmented by socio-economic status, there is a noticeable difference: respondents from lower socio-economic backgrounds were more likely to agree that the effectiveness of herbal remedies is due to the placebo effect (18.9%) compared to those from higher socio-economic backgrounds (8.7%). This disparity may reflect differences in scientific literacy, access to modern medical information, or cultural influences that affect beliefs about herbal medicine. Furthermore, the high percentage of disagreement across genders (over 53%) and age groups (ranging from 54% to 61%) suggests a broadly consistent belief in the genuine medicinal properties of herbal remedies across the community. These findings align with previous research indicating that although placebo effects may contribute to the perceived benefits of herbal treatments, the majority of users believe these remedies contain active compounds that produce real therapeutic effects [15] [16]. Nonetheless, the presence of a substantial minority accepting the placebo explanation underscores the need for ongoing public health education to enhance accurate understanding of how herbal medicines work. This can help individuals make informed choices about safe and effective treatment options.

### Willingness to Use *C. asiatica* for Treatment

Table 6 summarizes respondents’ attitudes toward the acceptability and perceived effectiveness of *Centella asiatica* (C.A.) in disease treatment, disaggregated by socio-economic status, sex, and age group. A vast minority of participants (91%) agreed that they would recommend *C. asiatica* to relatives and friends, and nearly 74% agreed that people would use *C. asiatica* (extract) if it was on the list of “western” medicines. These rates did not differ by areas, sex, or age group (χ^2^ test, p>0.05). About 39% of participants believed that *C. asiatica* was effective only if used in its natural form, 32.1% disagreed with this, and 28.1% were neutral. This percentage varied by area (χ^2^ test, p<0.05). Over 58% of people believed that a medicine derived from *C. asiatica* could be effective in treating liver cancer. However, significant differences emerge when examining responses across socio-economic strata.This share was higher among participants from high socio-economic areas than that of those from low socio-economic areas (63.2% vs 53%, χ^2^ test, p<0.001). Correspondingly, the proportion of respondents disagreeing with the efficacy of *Centella asiatica* in liver cancer treatment was more than double in the low socio-economic group (9.5%) compared to the high socio-economic group (4.2%).

**Table 6.** Acceptability of respondents in using *C. asiatica* for disease treatment.

| Indicator | Area |  | Sex |  | Age group |  |  | Overall<br>(n=804) |
| --- | --- | --- | --- | --- | --- | --- | --- | --- |
|  | Low socio-economic<br>(n=402), % | High socio-economic<br>(n=402), % | Male<br>(n=400), % | Female<br>(n=404), % | Aged under 40<br>(n=114), % | Aged 40-59<br>(n=291), % | Aged 60+<br>(n=399), % |  |
| I would recommend C.A to my family and friends |  |  |  |  |  |  |  |  |
| Disagree | 3.7 | 2.2 | 4.0 | 2.0 | 3.5 | 3.8 | 2.3 | 3.0 |
| Neutral | 5.2 | 6.7 | 6.3 | 5.7 | 7.0 | 7.2 | 4.7 | 6.0 |
| Agree | 91.1 | 91.1 | 89.7 | 92.3 | 89.5 | 89.0 | 93.0 | 91.0 |
| People would use C.A if it was put into 'Western' medicine |  |  |  |  |  |  |  |  |
| Disagree | 4.0 | 3.7 | 3.5 | 4.2 | 4.4 | 3.5 | 4.0 | 3.9 |
| Neutral | 21.6 | 23.4 | 25.0 | 20.1 | 28.1 | 19.9 | 22.8 | 22.5 |
| Agree | 74.4 | 72.9 | 71.5 | 75.7 | 67.5 | 76.6 | 73.2 | 73.6 |
| C.A is only effective if used naturally | * |  |  |  |  |  |  |  |
| Disagree | 34.6 | 29.6 | 35.0 | 29.2 | 43.9 | 33.3 | 27.8 | 32.1 |
| Neutral | 23.9 | 32.3 | 25.5 | 30.7 | 27.2 | 26.1 | 29.8 | 28.1 |
| Agree | 41.5 | 38.1 | 39.5 | 40.1 | 28.9 | 40.6 | 42.4 | 39.8 |
| C.A is effective in the treatment of liver cancer | *** |  |  |  |  |  |  |  |
| Disagree | 9.4 | 4.2 | 8.3 | 5.4 | 4.4 | 9.6 | 5.5 | 6.8 |
| Neutral | 37.6 | 32.6 | 31.7 | 38.4 | 34.2 | 33.0 | 36.9 | 35.1 |
| Agree | 53.0 | 63.2 | 60.0 | 56.2 | 61.4 | 57.4 | 57.6 | 58.1 |

Note: \*: $p<0.05$ , \*\*: $p<0.01$ and \*\*\*: $p<0.001$

This openness is significant because it highlights the potential for integrating traditional herbal remedies into more formal cancer care regimens, where *C. asiatica* could be used alongside chemotherapy, radiotherapy, or other modern cancer treatments. The public’s readiness to embrace such integration underscores the importance of further research into the medicinal benefits of *C. asiatica* in cancer therapy.

## Discussion

The results of this study provide valuable insights into the knowledge, attitudes, and beliefs of the Vietnamese population regarding the use of *C. asiatica* in the treatment of liver cancer. The widespread awareness of Rau Ma and its perceived benefits reflect a deeply rooted cultural trust in traditional medicine, alongside the increasing availability of modern medical treatments. The study also uncovers several important patterns and nuances that reveal significant socio-economic and regional differences in attitudes toward traditional and modern medicine.

### Traditional Medicine Use in the Context of Cancer Treatment

Vietnam has a long history of using traditional medicine, and in particular traditional medicine remains a preferred choice among rural ethnic minority communities [17]. In these regions, traditional remedies are often the first line of treatment, either as a complement to, or in place of, conventional medicine. The study found that a large proportion of respondents (91%) were willing to recommend *C. asiatica* for liver cancer treatment, indicating strong public confidence in traditional remedies. This highlights the Vietnamese public’s trust in herbal medicine, particularly for chronic diseases like cancer, where treatment options may seem out of reach, limited or ineffective [18][19]. Traditional medicine’s holistic approach, which focuses on overall well-being and health maintenance rather than simply targeting the disease, seems to resonate with many individuals, especially those in later stages of life or facing incurable conditions.

The high willingness to recommend *C. asiatica* also suggests that many people are not only familiar with traditional remedies but also view them as a legitimate and effective treatment option. This finding supports the idea that traditional medicine in Vietnam, particularly for cancer treatment, is not just a last resort but an important part of the overall healthcare framework. For many respondents, especially those in rural areas or lower socio-economic backgrounds, traditional remedies like *C. asiatica* offer a sense of control and hope in the face of challenging medical conditions [19] [20].

Moreover, the study found that respondents from lower socio-economic backgrounds exhibited a higher acceptance of *C. asiatica* as a complementary or alternative treatment. This trend is consistent with other studies in rural settings, where access to modern healthcare services may be more limited, and traditional remedies are often seen as more feasible and affordable. Traditional medicine, therefore, continues to play a central role in these communities, serving not only as a remedy for health conditions but also as a cultural anchor that connects individuals to their heritage and identity [16].

### Regional and Socio-Economic Variations

One of the most striking findings from this study is the regional and socio-economic variations in attitudes toward traditional and Western medicine. In higher socio-economic areas, where access to Western healthcare is more readily available, respondents showed a stronger preference for modern medical treatments [21]. These individuals were more likely to have knowledge of pharmaceutical drugs, more trust in their efficacy, and a lower confidence in traditional remedies like *C. asiatica*. This can be attributed to their greater exposure to scientifically tested treatments and healthcare systems that prioritize evidence-based approaches. For this group, traditional medicine is often viewed as a complementary rather than a standalone treatment option.

In contrast, respondents from higher socio-economic backgrounds, particularly those in rural areas, demonstrated higher acceptance of *C. asiatica* as a legitimate treatment for liver cancer. These respondents are often more reliant on affordable, accessible healthcare options, and traditional remedies provide a feasible alternative to expensive modern treatments. For many of these individuals, *C. asiatica* may serve as a vital support system when conventional treatments are either unavailable or unaffordable [22]. This highlights a socio-economic divide in healthcare access, where lower-income individuals are more likely to turn to traditional remedies due to their lower cost and greater accessibility.

### Knowledge and Education About Traditional Medicine

While 87.3% of participants reported familiarity with *C. asiatica*, noticeable differences emerged in the depth of knowledge regarding its medicinal properties across socio-economic groups. As anticipated, individuals from higher socio-economic backgrounds demonstrated more comprehensive understanding about the herb’s therapeutic benefits, reflecting greater access to healthcare information, education, and exposure to scientific research on traditional medicines [23]. This finding aligns with expectations that knowledge transmission would be more thorough in these groups.

Surprisingly, however, a substantial proportion of respondents from lower socio-economic groups also recognized the medicinal value of *C. asiatica*, despite possessing less detailed knowledge of its pharmacological properties [24]. This suggests that, beyond formal channels, traditional knowledge is effectively transmitted through informal networks within families and communities. This unexpected diffusion underscores the resilience and cultural embedding of traditional medicinal knowledge, even among populations with limited access to formal education and scientific resources.

The findings also pointed to a lack of understanding regarding the clinical evidence supporting the use of *C. asiatica* in cancer treatment. Many participants expressed positive beliefs about its efficacy, but these beliefs were not necessarily grounded in scientific evidence. This highlights a critical gap in the education and awareness surrounding scientifically backed research on traditional remedies. Public health campaigns could play a vital role in educating the public about the clinical research on herbal treatments like *C. asiatica*, thereby promoting a better understanding of their potential and limitations [25].

There is also a strong conception that traditional and herbal medicines are ‘safer’. However, there is a need to be careful as this can be a misconception for ‘cross-over’ drugs containing derivatives from well recognised traditional remedies, which are no longer ‘natural’, or share such characteristics with the plant from which they are extracted. Although this idea could enhance acceptance of cross overs, this requires careful discussion and engagement to ensure the changing form of ‘cross over drugs’ and their relation to their origin plants in well understood by patients and communities.

### Potential for Integrating Traditional Medicine with Western Healthcare

A key finding from the study is the openness of participants to using traditional and herbal medicines, and specifically *C. asiatica*, in combination with Western treatments for liver cancer. 74% of respondents indicated that they would be willing to incorporate traditional remedies into their treatment regimen if they were integrated into conventional Western medical plans. This suggests that there is a recognition of the complementary role that traditional medicine can play alongside modern medical approaches, particularly in managing chronic illnesses like cancer [4].

This hybrid approach to treatment is becoming increasingly popular worldwide, as patients seek complementary therapies to help manage the side effects of conventional treatments like chemotherapy and radiotherapy. In Vietnam, where both traditional and modern healthcare systems coexist, there is a unique opportunity to combine the strengths of both. For instance, *C. asiatica* has shown promising anti-inflammatory and anti-cancer properties in preliminary studies, which could help alleviate some of the debilitating side effects of cancer treatment, such as fatigue, weakness, and digestive issues [4–6].

Integrating *C. asiatica* into cancer care regimens could provide holistic benefits by improving the overall quality of life for patients, especially in terms of symptom management. However, it is important to acknowledge that the efficacy and safety of herbal medicines like *C. asiatica* can be significantly influenced by quality control factors. Variations in cultivation conditions, geographic origin, and processing methods may lead to substantial differences in the levels of active compounds, as demonstrated in recent studies on *C. asiatica* [26]. These inconsistencies highlight the need for rigorous standardization and quality assurance to ensure consistent therapeutic outcomes. Moreover, further well-designed clinical trials are essential to substantiate the potential benefits and safety of *C. asiatica* in cancer treatment, especially when used concurrently with conventional therapies. Establishing comprehensive regulatory frameworks is critical to govern the safe and appropriate use of traditional medicines. Such regulations should address quality control, dosage standardization, and potential herb-drug interactions to minimize risks and maximize patient safety.

### Limitations and Future Research Directions

While this study provides important insights, there are some limitations that should be considered. First, the survey relied on self-reported data, which can be subject to biases, especially when participants overestimate their knowledge or experience with traditional remedies. Additionally, the study did not explore the clinical efficacy of *C. asiatica* in cancer treatment, and further rigorous clinical trials are needed to validate its therapeutic potential.

During the survey, only 80 out of the initially targeted 100 households in Quang Tho commune, Quang Dien district (Hue city), were successfully interviewed, as many residents were absent due to offshore fishing activities. To achieve the required sample size, an additional 20 households from Quang Phu commune, aan adjacent locality within the same district.

Although, the original sampling strategy stratified participants by two age group (under 60 and 60 years and above), subsequent analysis expanded the categorization into three age groups—young (under 40), middle-aged (40–59), and older adults (60 and above). This reclassification lead to an uneven distribution across agre groups. Nonetheless,, the sample remains valid and statistically robust for the intended analysis

Recruiting respondents for studies involving cancer patients and their caregivers presented several challenges, particularly when focusing on traditional herbal medicine such as *Centella asiatica* remedies. One significant difficulty lied in the willingness of individuals to participate; patients and families coping with cancer often experience emotional distress and fatigue, making them less inclined to engage in research activities. Additionally, approaching families with members affected by cancer required sensitivity and trust-building, as these conversations could be perceived as intrusive or overwhelming. Accessing a representative sample was further complicated by the limited awareness and use of traditional herbal treatments like C.A., which may not be widely recognized or accepted in some communities. This lack of familiarity restricted the pool of potential respondents knowledgeable about or willing to discuss these remedies, thus impacting recruitment and the generalizability of study findings.

Future research could focus on randomized controlled trials (RCTs) to examine the effectiveness of *C. asiatica* in treating liver cancer and other types of cancer. Furthermore, research into its pharmacological properties and how it interacts with conventional cancer treatments is essential. Understanding its dosage and potential contraindications will be key to its safe integration into cancer care regimens.

Public health research should also focus on evaluating the impact of educational campaigns aimed at improving the public’s understanding of both traditional and modern treatments. Longitudinal studies could explore the long-term effects of integrating traditional medicine with Western cancer therapies, providing valuable data on its safety and efficacy.

## Conclusion

The study underlines the significant role of traditional medicine, particularly *Centella asiatica* (Rau Ma), continues to play in the treatment landscape of chronic conditions such as liver cancer in Vietnam. The widespread familiarity and acceptance of *C. asiatica* across diverse socio-economic and regional groups underscore its deep cultural integration and potential as a complementary therapy alongside conventional cancer treatments. Notably, regional and socio-economic disparities shape perceptions and usage patterns, with rural and lower socio-economic populations exhibiting stronger reliance on traditional remedies due to accessibility and affordability considerations.

While public openness toward integrating *C. asiatica* into modern cancer care is promising, the study also identifies gaps in scientific evidence and awareness regarding its efficacy and safety. This underscores the urgent need for rigorously designed clinical trials to substantiate therapeutic benefits and to establish clear pharmacological profiles, including dosage, potential interactions, and contraindications. Furthermore, quality control remains a vital concern given the variability in active compound concentrations stemming from differences in cultivation and processing practices..

To fully realize the potential of *C. asiatica* within integrative oncology, a comprehensive framework that combines clinical research, pharmacological investigation, and regulatory oversight is essential. Public health initiatives should also aim to enhance patient and caregiver education, promoting informed decision-making regarding the use of traditional herbal medicines in cancer care. Collectively, these efforts can help bridge the gap between traditional knowledge and evidence-based medicine, advancing holistic and accessible cancer treatment options for the Vietnamese population.

## Data Availability

All relevant data are within the manuscript and its Supporting Information files.

## Supporting information

**S1 Table. Data analysis**

(XLSX)

**S2 File. Survey Tools (Questionnaire, Terms, Enumerator assessment, Consent form)**

(DOCX)

## Acknowledgements

We would like to express our gratitude to the participants in Hanoi and Hue for their valuable time and responses. Our sincere thanks also go to the remunerators who assisted us in collecting data, as well as to our colleagues from the UV-CAP project, partners from Quang Tho II Cooperative, the National Cancer Institute, and the Commune People’s Committees of the target communes for their support and providing the necessary conditions to carry out this study. We thank the UK’s Engineering and Physical Science Research Council and Global Challenges Research Fund (EP/T020164/1) for financial support.

## Author Contributions

**Conceptualization:**Rebeca Casidy; Yen Be Thi Hoang

**Data curation:** Ky The Hoang

**Funding acquisition:** Christopher J. Serpell

**Methodology:** Yen Be Thi Hoang, Ky The Hoang, Rebeca Casidy

**Project administration:** Yen Be Thi Hoang, Christopher J. Serpell

**Software:** Ky The Hoang

**Supervision**: Dinh Quoc Huy

**Validation:** Yen Be Thi Hoang, Ky The Hoang

**Writing – original draft:** Yen Be Thi Hoang

**Writing – review & editing:** Ky The Hoang, Rebeca Casidy, Christopher J. Serpell, Michelle D. Garrett

